# Hypoperfusion Profiles in outcome after Endovascular Therapy for Isolated Internal Carotid Artery Occluded Patients

**DOI:** 10.1101/2024.09.18.24313935

**Authors:** Feina Shi, Yigang Chen, Xu Zheng, Yun Jiang, Ziqi Xu, En Wang, Changzhu Wu, Chuan Xu, Qingqing Gao, Xi Hu, Jinhua Zhang

**Affiliations:** Department of Neurology, Sir Run Run Shaw Hospital of Zhejiang University, School of Medicine, Hangzhou, China; Department of Neurology, the First Affiliated Hospital of Zhejiang University, School of Medicine, Hangzhou, China; Department of Neurology, Taizhou Hospital of Zhejiang Province, Taizhou, China; Department of Radiology, Sir Run Run Shaw Hospital of Zhejiang University, School of Medicine, Hangzhou, China

**Author notes:** **Corresponding author**: Jinhua Zhang M.D. Department of Neurology, Sir Run Run Shaw Hospital, Zhejiang University School of Medicine Hangzhou, Zhejiang, China. 310009.

**Keywords:** acute ischemic stroke, symptomatic isolated internal carotid artery occlusion, T_max_ mismatch ratio, neurological deterioration, outcome

## Abstract

**Background:** Patients with acute symptomatic isolated internal carotid artery occlusions (iICAO), also known as ICA-I occlusion, frequently presented with minor neurological deficits. However, they remained at risk for subsequent neurological deterioration even after intravenous thrombolysis. The efficacy of endovascular treatment (EVT) in iICAO patients remains uncertain. Recently, some retrospective studies revealed EVT for acute iICAO was safe and might confer potential benefits. Our aim was to identify the hypoperfusion profiles associated with neurological deterioration and functional outcome following EVT in iICAO patients.

**Methods:** We performed a multi-center, real-world cohort study involving acute symptomatic iICAO patients who underwent EVT from August 2018 to February 2024 across 3 stroke centers. Hypoperfusion profiles were assessed using T_max_ volumes (>4s, >6s, >8s, and >10s) and T_max_ mismatch ratios (>10s/>8s, >10s/>6s, >10s>4s, >8s/>6s, >8s/>4s, and >6s/>4s). Neurological deterioration was defined as an increase of ⩾4 points in the NIHSS score prior to EVT. Good outcome was defined as modified Rankin Score 0 - 1 at 90 days.

**Results:** A total of 63 patients were included in the final analysis. Multivariable models showed that a lower T_max_ ratio of >6s/>4s was an independent predictor for increased risk of neurological deterioration (OR, 0.003; 95% CI: 0.000-0.133; P = 0.003) and was associated with a better outcome following EVT (OR, 0.066; 95% CI: 0.005-0.874; P = 0.039) after adjustment. The optimal threshold for the T_max_ >6s/>4s ratio was identified as 0.5.

**Conclusions:** A perfusion defect severity ratio of T_max_ >6s/>4s less than 0.5 might indicate the likelihood of neurological deterioration, yet it would be associated with achieving a good outcome following EVT for iICAO patients. The hypoperfusion profile characterized by a T_max_ >6s/>4s might be useful to the neurointerventionists for facilitating appropriate patient triage and guiding prognosis for EVT in iICAO patients.

## Introduction

Isolated internal carotid artery occlusion (iICAO), without associated occlusions of the circle of Willis (ie, without T-shaped, L-shaped, or tandem occlusions), was not rare among acute ischemic stroke (AIS) patients, accounting for approximately 10%-20% of all carotid occlusions^1–3^. Although many clinical trials have suggested the benefit of endovascular treatment (EVT) for AIS patients with intracranial large vessel occlusion (LVO) ^4,5^, those patients with iICAO have often been overlooked. Given the lack of evidence derived from randomized trials, the potential benefits of EVT for the AIS patients only with iICAO remains unclear. Recently, several non-randomized studies have demonstrated the feasibility, safety, and potential benefits of EVT for iICAO patients^6,7^.

Clinical severity among patients with iICAO is highly variable and they frequently present with minor neurological deficits in part as a function of collateral circulation^8^. The iICAO patients often exhibit extensive hypoperfusion on imaging, indicating insufficient cerebral perfusion despite good arterial collateral circulation. The cerebral collateral cascade, facilitating the successful delivery of blood flow to ischemic tissue, would influence the clinical and radiological outcomes in AIS patients ^9^. For iICAO patients allowing blood flow to the middle cerebral artery through the circle of Willis, the arterial collateral circulation on CT angiography (CTA) seemed adequate. However, the extensive hypoperfusion observed in iICAO patients still indicates inadequacy of collateral supply. Thus, the extensive hypoperfusion observed in iICAO patients might be the result of the complex vascular mechanisms, rather than solely by arterial collaterals. It has been demonstrated that perfusion delay lesions would be related with neurological deterioration^10^. In previous studies, early neurological deterioration was reported in 15-30% of these iICAO patients even after intravenous thrombolysis or when presenting with mild symptoms initially ^8,11^. Therefore, it is crucial to identify iICAO patients who are at high risk for neurological deterioration, as they are the individuals most likely to benefit from these promising therapies, such as EVT. CT perfusion (CTP) is increasingly applied to quantitative evaluation of perfusion status of the whole brain in AIS patients. In clinical trials, CTP has been used to select optimal candidates who would benefit from reperfusion therapy. The characteristics of hypoperfused brain regions and their relationship with outcome after EVT for iICAO patients still remain unclear. In previous studies, hypoperfusion profiles evaluated with time-to-peak of the residual function (T_max_) ratios were found to be associated with infarct growth and functional outcome^12–14^.

In this study, we aimed to explore hypoperfusion profiles of iICAO associated with neurological deterioration. We also evaluated the relationship between hypoperfusion profiles and functional outcome after EVT in iICAO patients.

## Material and Methods

### Ethics Statement

The investigations have been conducted according to the principles expressed in the Declaration of Helsinki. The protocol was approved by the local human ethics committees. Written informed consent was obtained from all patients or their designated proxy to confirm the acceptance of reperfusion therapy.

### Patients

The current multi-center real-world retrospective cohort study reviewed acute ischemic stroke patients who were treated with EVT from August 2018 to February 2024 from 3 stroke centers. Then, patients were included if they (1) had a recent diagnosis (<2 weeks) of AIS confirmed with by DWI or CT; (2) underwent CTP before EVT treatment; and (3) had ipsilateral ICAO without occlusion of the ipsilateral intracranial large arteries, such as C7 segment of ICA, M1–M2 segments of the middle cerebral artery, A1-A2 segments of the anterior cerebral artery, P1-P2 segments of the posterior cerebral artery. Patients who had poor image quality due to motion artifacts or incomplete images were excluded.

### Clinical data

Baseline clinical variables and radiologic data were recorded, including demographics, risk factors (smoking, hypertension, diabetes mellitus, hyperlipidaemia, a history of stroke/TIA and atrial fibrillation), prior usage of antiplatelet/ anticoagulants, intravenous thrombolysis, onset-to-puncture time (OPT), baseline National Institutes of Health Stroke Scale (NIHSS) score. The stroke etiologies were determined as large-artery atherosclerosis (LAA), cardioembolism (CE), dissection, other causes or undetermined^7^. Neurological deterioration was defined as an increase of ⩾ 4 points in the NIHSS score before EVT^15^. Patients were then dichotomized into good outcome (mRS score of ≤1) and poor outcome (mRS score of >1) at 90 days.

### Imaging Analysis

The occlusion location was determined on reconstructed CTA images derived from CTP. Recanalization after EVT was assessed on DSA images as modified TICI grades ≥2b, and hemorrhagic transformation (HT) was assessed on follow-up NCCT according to the second European-Australasian Acute Stroke Study (ECASS Ⅱ)^16^. Pretreatment relative cerebral blood flow (CBF) of less than 30% at CTP was used to calculate the baseline infarct core volume, and T*_max_* was used for volumetric measurement of pretreatment hypoperfusion lesion profiles. The hypoperfused brain tissue volumes were automatically measured using T_max_ thresholds of >10s, >8s, >6s, and >4s. T_max_ ratios of >10s/>8s, >10s/>6s, >10s>4s, >8s/>6s, >8s/>4s, and >6s/>4s were considered T_max_ mismatch ratios. All imaging analyses were assessed by the independent experienced neuroradiologist.

### Statistical Analysis

Continuous variables were reported as medians with interquartile range, and categorical variables were reported as proportions. For categorical variables, Chi-Square test was used to compare differences among groups. For continuous variables, Man-Whitney U test was used to compare the difference between two groups. The receiver operating characteristic (ROC) analyses were performed to assess the discriminative ability and area under the curve (AUC) was calculated. Optimal cutoff values were derived from ROC curves, and sensitivity and specificity were calculated on the basis of these best cutoff values. The variables with P values <0.1 at univariable analysis were included in the multivariable models. Multivariable regression analysis was performed using the method of backward. All statistical analyses were performed using SPSS, Version 22.0 (IBM, Armonk, New York). P value < 0.05 was considered statistically significant.

## Results

### Patient Characteristics

A total of 65 patients met the inclusion criteria and 2 patients were excluded due to poor image quality. Finally, a total of 63 patients with AIS attributable to iICAO were included in final analysis, with a median age of 66 (55-72) years, a median NIHSS score of 9 (6-14), and a median OPT of 993 (660-2001) min. A median of baseline infarct core (CBF <30%) volume was 0.9 (0.0-14.7) mL, and a median of T_max_>6s volume was 142.6 (52.8-234.0) mL. Intravenous thrombolysis was administered to 7 (11.1%) patients. Neurological deterioration occurred prior to EVT in 34 (54.0%) iICAO patients. Among these, 59 (93.7%) iICAO patients achieved successful recanalization, and hemorrhagic transformation was observed in 11(17.5%) iICAO patients after EVT.

### Association of hypoperfusion profiles with neurological deterioration in iICAO patients

As show in Table 1, the iICAO patients with neurological deterioration had a significantly lower NIHSS score (median 7 vs 11, P<0.001) and a lower ASPECTS score (median 8 vs 9, P=0.023) than those without neurological deterioration. The iICAO patients with neurological deterioration had a significantly lower baseline infarct core volume (median 0.0mL vs 8.3mL, P=0.006). Compared to those without neurological deterioration, iICAO patients with neurological deterioration exhibited significantly lower volumes of T_max_>6s (median 105.7mL vs 197.4mL, P=0.034), T_max_>8s (median 32.1mL vs 111.0mL, P=0.021), T_max_>10s (median 6.4mL vs 40.7 mL, P=0.003). However, there was no significant association between T_max_>4s volume and neurological deterioration. Furthermore, T_max_ mismatch ratios, including T_max_ ratios of >10s/8s (median 0.3 vs 0.6, P<0.001), >10s/>6s (median 0.1 vs 0.3, P=0.001), >10s/>4s (median 0.0 vs 0.1, P=0.001), >8s/>6s (median 0.4 vs 0.6, P=0.004), >8s/>4s (median 0.1 vs 0.3, P=0.003), >6s/>4s (median 0.4 vs 0.6, P=0.001), were significantly lower in iICAO patients with neurological deterioration compared to those without neurological deterioration.

**Table 1.**
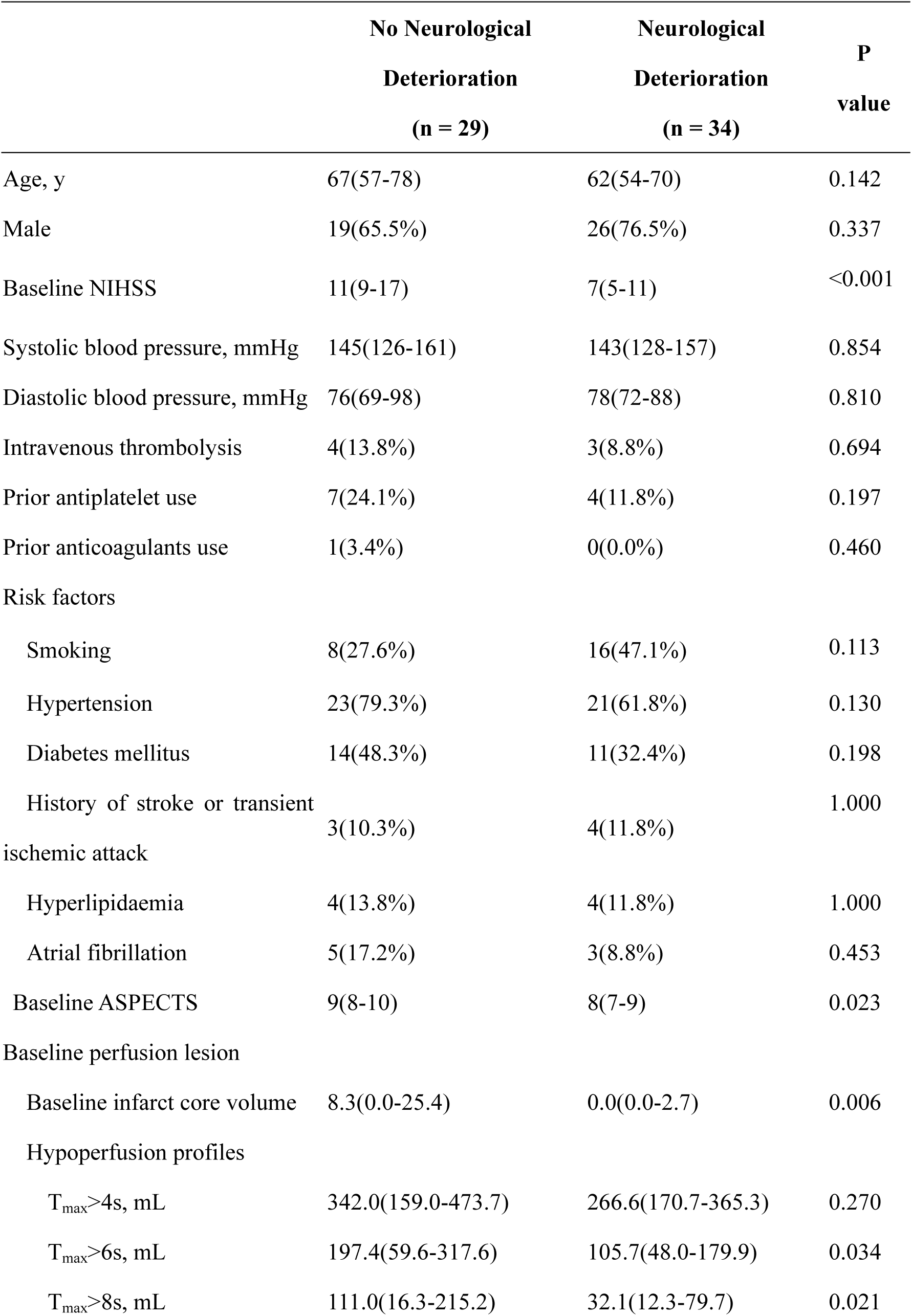
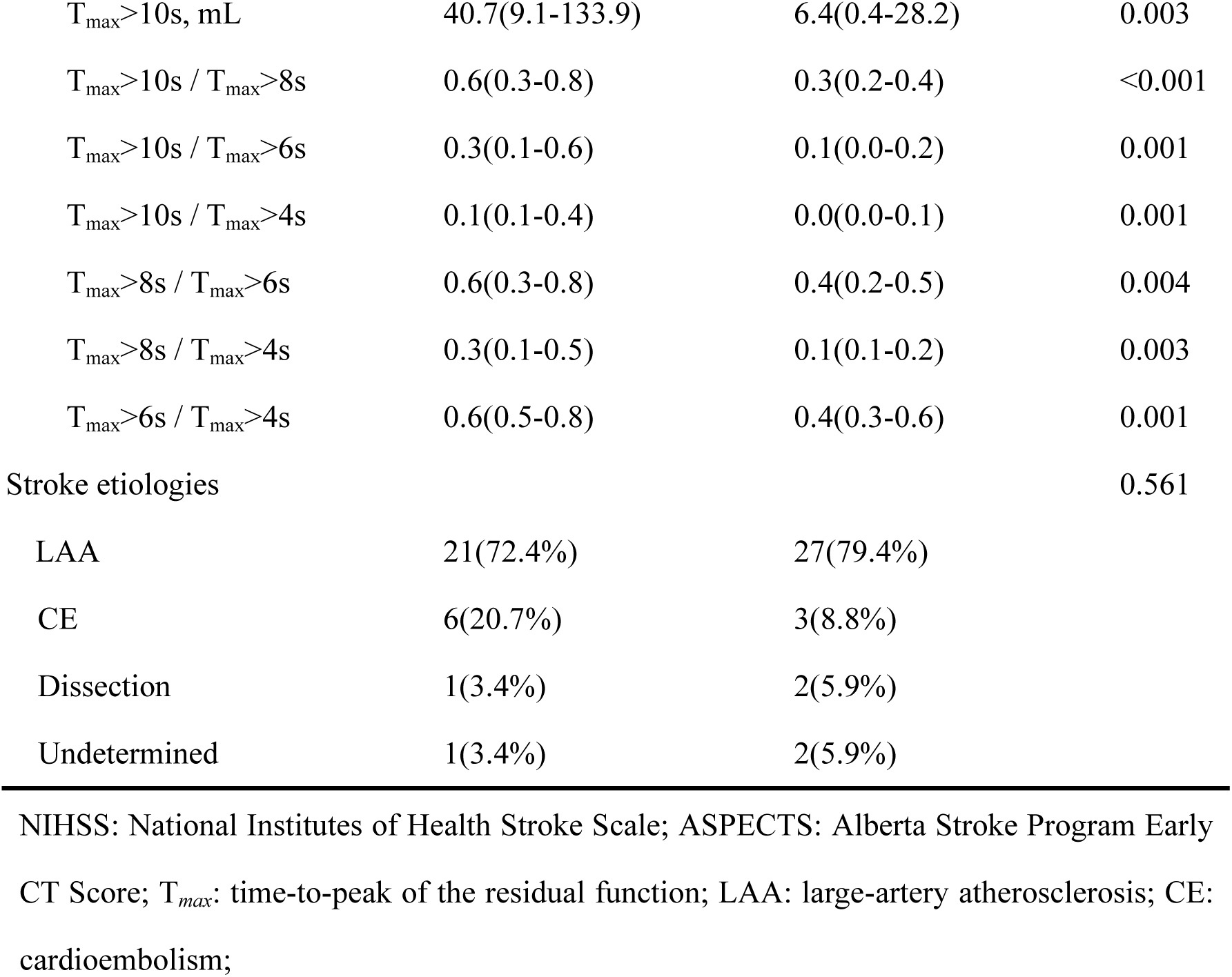
Comparison of Characteristics between Symptomatic Isolated Internal Carotid Artery Occlusion Patients with Neurological Deterioration.

|  | No Neurological<br>Deterioration<br>(n = 29) | Neurological<br>Deterioration<br>(n = 34) | P<br>value |
| --- | --- | --- | --- |
| Age, y | 67(57-78) | 62(54-70) | 0.142 |
| Male | 19(65.5%) | 26(76.5%) | 0.337 |
| Baseline NIHSS | 11(9-17) | 7(5-11) | <0.001 |
| Systolic blood pressure, mmHg | 145(126-161) | 143(128-157) | 0.854 |
| Diastolic blood pressure, mmHg | 76(69-98) | 78(72-88) | 0.810 |
| Intravenous thrombolysis | 4(13.8%) | 3(8.8%) | 0.694 |
| Prior antiplatelet use | 7(24.1%) | 4(11.8%) | 0.197 |
| Prior anticoagulants use | 1(3.4%) | 0(0.0%) | 0.460 |
| Risk factors |  |  |  |
| Smoking | 8(27.6%) | 16(47.1%) | 0.113 |
| Hypertension | 23(79.3%) | 21(61.8%) | 0.130 |
| Diabetes mellitus | 14(48.3%) | 11(32.4%) | 0.198 |
| History of stroke or transient<br>ischemic attack | 3(10.3%) | 4(11.8%) | 1.000 |
| Hyperlipidaemia | 4(13.8%) | 4(11.8%) | 1.000 |
| Atrial fibrillation | 5(17.2%) | 3(8.8%) | 0.453 |
| Baseline ASPECTS | 9(8-10) | 8(7-9) | 0.023 |
| Baseline perfusion lesion |  |  |  |
| Baseline infarct core volume | 8.3(0.0-25.4) | 0.0(0.0-2.7) | 0.006 |
| Hypoperfusion profiles |  |  |  |
| T<sub>max</sub>>4s, mL | 342.0(159.0-473.7) | 266.6(170.7-365.3) | 0.270 |
| T<sub>max</sub>>6s, mL | 197.4(59.6-317.6) | 105.7(48.0-179.9) | 0.034 |
| T<sub>max</sub>>8s, mL | 111.0(16.3-215.2) | 32.1(12.3-79.7) | 0.021 |
| $T_{\max} > 10s$, mL | 40.7(9.1-133.9) | 6.4(0.4-28.2) | 0.003 |
| $T_{\max} > 10s$ / $T_{\max} > 8s$ | 0.6(0.3-0.8) | 0.3(0.2-0.4) | <0.001 |
| $T_{\max} > 10s$ / $T_{\max} > 6s$ | 0.3(0.1-0.6) | 0.1(0.0-0.2) | 0.001 |
| $T_{\max} > 10s$ / $T_{\max} > 4s$ | 0.1(0.1-0.4) | 0.0(0.0-0.1) | 0.001 |
| $T_{\max} > 8s$ / $T_{\max} > 6s$ | 0.6(0.3-0.8) | 0.4(0.2-0.5) | 0.004 |
| $T_{\max} > 8s$ / $T_{\max} > 4s$ | 0.3(0.1-0.5) | 0.1(0.1-0.2) | 0.003 |
| $T_{\max} > 6s$ / $T_{\max} > 4s$ | 0.6(0.5-0.8) | 0.4(0.3-0.6) | 0.001 |
| Stroke etiologies |  |  | 0.561 |
| LAA | 21(72.4%) | 27(79.4%) |  |
| CE | 6(20.7%) | 3(8.8%) |  |
| Dissection | 1(3.4%) | 2(5.9%) |  |
| Undetermined | 1(3.4%) | 2(5.9%) |  |

NIHSS: National Institutes of Health Stroke Scale; ASPECTS: Alberta Stroke Program Early CT Score; $T_{\max}$ : time-to-peak of the residual function; LAA: large-artery atherosclerosis; CE: cardioembolism;

Then, the baseline NIHSS score, ASPECTS score, baseline infarct core volume, and alongside with each hypoperfusion profile with P < 0.1 from the univariable analyses (i.e., the volume of T_max_>6s, T_max_>8s, T_max_>10s, and T_max_ ratios of >10s/8s, >10s/6s, >10s/>4s, >8s/6s, >8s/>4s, >6s/>4s), were included in a separate multivariable regression analysis. Multivariable models showed that T_max_ ratio of >6s/>4s (OR, 0.003; 95% CI: 0.000-0.133; P = 0.003), T_max_ ratio of >8s/>4s (OR, 0.010; 95% CI: 0.000-0.510; P = 0.022), and the volume of T_max_>6s (OR, 0.994; 95% CI: 0.987-1.000; P = 0.049) were all independently associated with neurological deterioration after adjustment, respectively. The other hypoperfusion profiles did not emerge as independent factors of neurological deterioration (all P>0.05). The ROC analyses indicated that the optimal cutoff, AUC, sensitivity, specificity and Youden index for T_max_ ratio of >6s/>4s in predicting neurological deterioration were 0.5, 79.3%, 70.6% and 49.9%, respectively. For T_max_ ratio of >8s/>4s, these values were 0.2, 65.5%, 76.5%, and 42.0%. And these values were 164.2mL, 65.5%, 73.5%, and 39.0% for T_max_>6s (Table 2).

**Table 2.** Ability of Perfusion Lesion to Discriminate Neurological Deterioration in Patients with Symptomatic Isolated Internal Carotid Artery Occlusion before Endovascular Thrombectomy

| Variables | AUC (95% CI) | Cut off | Sensitivity | Specificity | Youden index | P value |
| --- | --- | --- | --- | --- | --- | --- |
| Baseline infarct core volume | 0.691(0.555-0.826) | 2.9mL | 62.1% | 79.4% | 41.5% | 0.006 |
| Hypoperfusion profiles |  |  |  |  |  |  |
| T<sub>max</sub>>4s, mL | 0.581(0.434-0.728) | 424.5mL | 31.0% | 91.2% | 22.2% | 0.278 |
| T<sub>max</sub>>6s, mL | 0.656(0.514-0.798) | 164.2mL | 65.5% | 73.5% | 39.0% | 0.031 |
| T<sub>max</sub>>8s, mL | 0.670(0.532-0.809) | 46.7mL | 69.0% | 67.6% | 36.6% | 0.016 |
| T<sub>max</sub>>10s, mL | 0.716(0.587-0.845) | 27.0mL | 62.1% | 76.5% | 38.6% | 0.001 |
| T<sub>max</sub>>10s / T<sub>max</sub>>8s | 0.762(0.640-0.883) | 0.3 | 89.7% | 56.3% | 46.0% | <0.001 |
| T<sub>max</sub>>10s / T<sub>max</sub>>6s | 0.753(0.632-0.875) | 0.2 | 72.4% | 72.7% | 45.1% | <0.001 |
| T<sub>max</sub>>10s / T<sub>max</sub>>4s | 0.751(0.628-0.873) | 0.1 | 79.3% | 64.7% | 44.0% | <0.001 |
| T<sub>max</sub>>8s / T<sub>max</sub>>6s | 0.713(0.583-0.842) | 0.4 | 69.0% | 66.7% | 35.7% | 0.001 |
| T<sub>max</sub>>8s / T<sub>max</sub>>4s | 0.720(0.628-0.873) | 0.2 | 65.5% | 76.5% | 42.0% | 0.001 |
| T<sub>max</sub>>6s / T<sub>max</sub>>4s | 0.751(0.624-0.877) | 0.5 | 79.3% | 70.6% | 49.9% | <0.001 |

T<sub>max</sub>: time-to-peak of the residual function; AUC: area under the curve.

### Association of hypoperfusion profiles with functional outcome in iICAO patients receiving EVT

Among 57 iICAO patients with pre-stroke mRS <1, good outcome (i.e., 90-day mRS <=1) after EVT was achieved in 23(40.4%) patients. As illustrated in Table 3, the iICAO patients with good outcome had a significantly lower NIHSS score (median 8 vs 11, P=0.010). T_max_ volumes at T_max_>6s (median 55.7mL vs 164.2mL, P=0.006), T_max_>8s (median 14.7mL vs 66.4mL, P=0.007), and T_max_>10s (median 3.3mL vs 19.9mL, P=0.009) were significantly lower in iICAO patients with good outcome than those with poor outcome. The volume of T_max_>4s was inclined to be lower in iICAO patients with good outcome compared to those with poor outcome (median 189.3mL vs 309.0mL, P=0.053). T_max_ mismatch ratios of >10s/>4s (median 0.0 vs 0.1, P=0.034), >8s/>4s (median 0.1 vs 0.2, P=0.033), >6s/>4s (median 0.4vs 0.5, P=0.037) were also significantly lower in iICAO patients with good outcome than those with poor outcome. There were no significant differences between these two groups in baseline infarct core volume, nor in the other hypoperfusion profiles (T_max_ ratios of >10s/8s, >10s/>6s, >8s/>6s). We found that neurological deterioration was not related with the functional outcome after EVT in iICAO patients. There were no significant differences in recanalization rates, hemorrhagic transformation following EVT, or stroke etiologies between patients with good outcome and poor outcome.

**Table 3.**
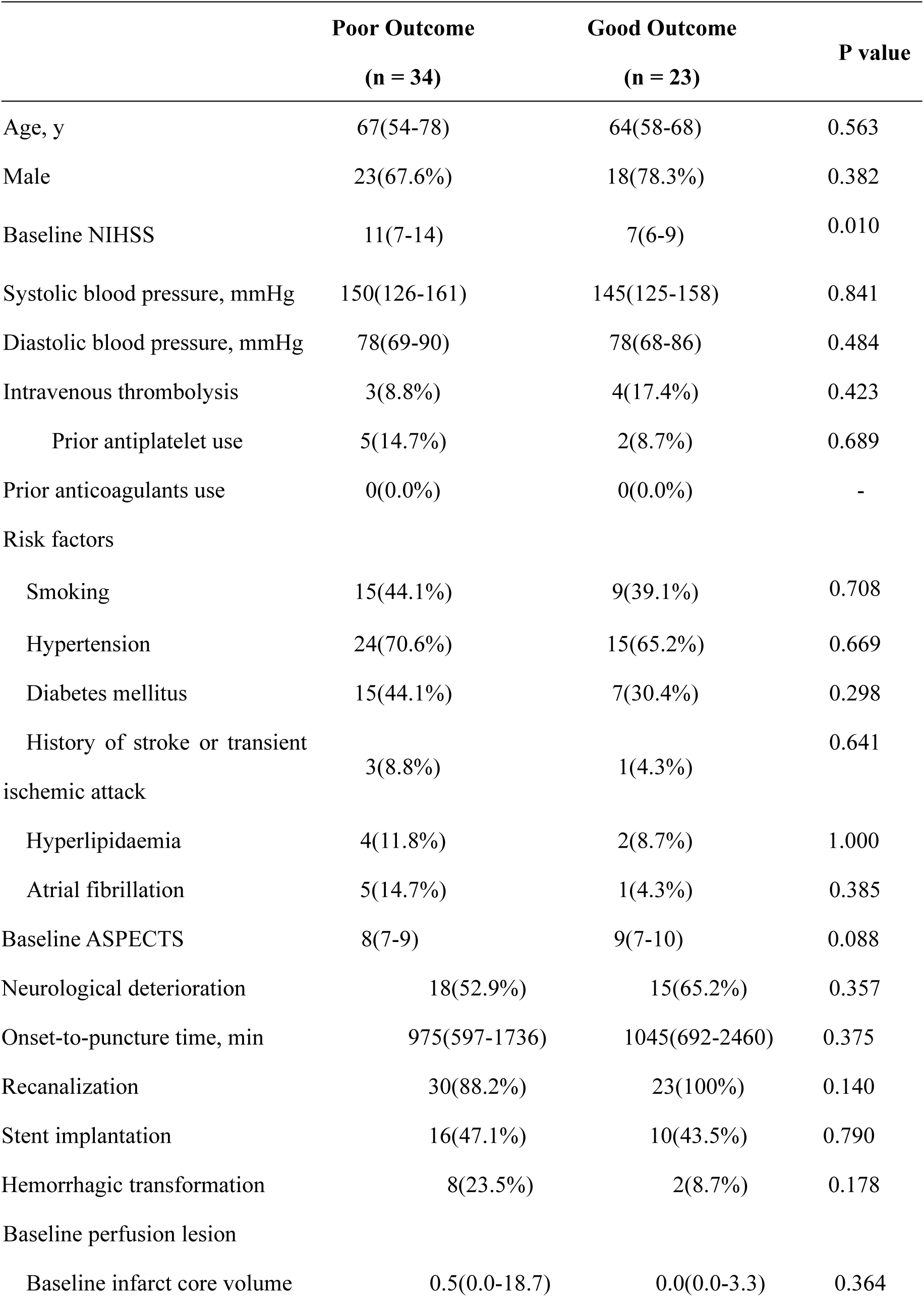
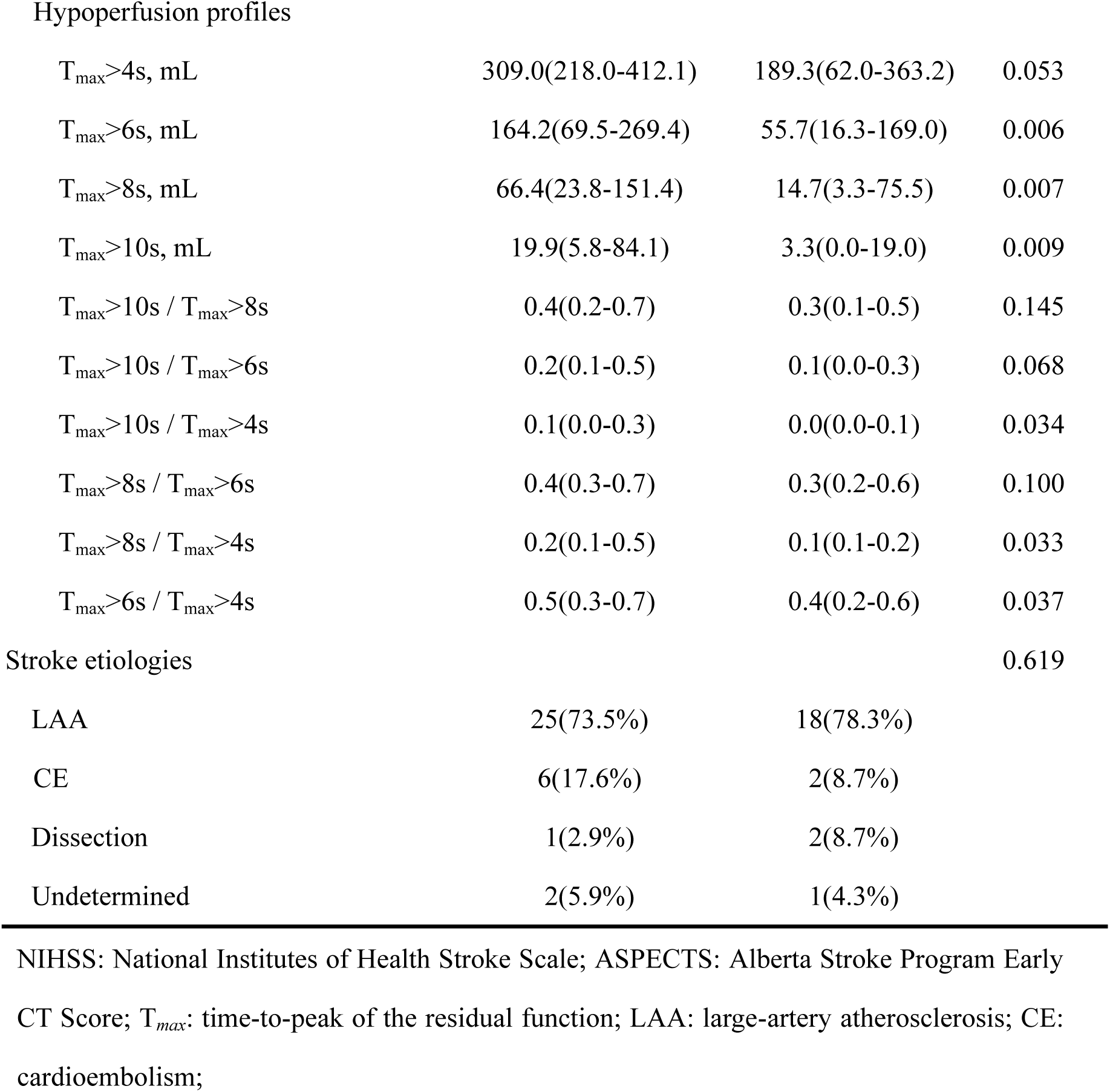
Comparison of Characteristics between Symptomatic Isolated Internal Carotid Artery Occlusion Patients Undergoing Endovascular Treatment with Good Outcome and Poor outcome

|  | Poor Outcome<br>(n = 34) | Good Outcome<br>(n = 23) | P value |
| --- | --- | --- | --- |
| Age, y | 67(54-78) | 64(58-68) | 0.563 |
| Male | 23(67.6%) | 18(78.3%) | 0.382 |
| Baseline NIHSS | 11(7-14) | 7(6-9) | 0.010 |
| Systolic blood pressure, mmHg | 150(126-161) | 145(125-158) | 0.841 |
| Diastolic blood pressure, mmHg | 78(69-90) | 78(68-86) | 0.484 |
| Intravenous thrombolysis | 3(8.8%) | 4(17.4%) | 0.423 |
| Prior antiplatelet use | 5(14.7%) | 2(8.7%) | 0.689 |
| Prior anticoagulants use | 0(0.0%) | 0(0.0%) | - |
| Risk factors |  |  |  |
| Smoking | 15(44.1%) | 9(39.1%) | 0.708 |
| Hypertension | 24(70.6%) | 15(65.2%) | 0.669 |
| Diabetes mellitus | 15(44.1%) | 7(30.4%) | 0.298 |
| History of stroke or transient<br>ischemic attack | 3(8.8%) | 1(4.3%) | 0.641 |
| Hyperlipidaemia | 4(11.8%) | 2(8.7%) | 1.000 |
| Atrial fibrillation | 5(14.7%) | 1(4.3%) | 0.385 |
| Baseline ASPECTS | 8(7-9) | 9(7-10) | 0.088 |
| Neurological deterioration | 18(52.9%) | 15(65.2%) | 0.357 |
| Onset-to-puncture time, min | 975(597-1736) | 1045(692-2460) | 0.375 |
| Recanalization | 30(88.2%) | 23(100%) | 0.140 |
| Stent implantation | 16(47.1%) | 10(43.5%) | 0.790 |
| Hemorrhagic transformation | 8(23.5%) | 2(8.7%) | 0.178 |
| Baseline perfusion lesion |  |  |  |
| Baseline infarct core volume | 0.5(0.0-18.7) | 0.0(0.0-3.3) | 0.364 |
| Hypoperfusion profiles |  |  |  |
| T<sub>max</sub>>4s, mL | 309.0(218.0-412.1) | 189.3(62.0-363.2) | 0.053 |
| T<sub>max</sub>>6s, mL | 164.2(69.5-269.4) | 55.7(16.3-169.0) | 0.006 |
| T<sub>max</sub>>8s, mL | 66.4(23.8-151.4) | 14.7(3.3-75.5) | 0.007 |
| T<sub>max</sub>>10s, mL | 19.9(5.8-84.1) | 3.3(0.0-19.0) | 0.009 |
| T<sub>max</sub>>10s / T<sub>max</sub>>8s | 0.4(0.2-0.7) | 0.3(0.1-0.5) | 0.145 |
| T<sub>max</sub>>10s / T<sub>max</sub>>6s | 0.2(0.1-0.5) | 0.1(0.0-0.3) | 0.068 |
| T<sub>max</sub>>10s / T<sub>max</sub>>4s | 0.1(0.0-0.3) | 0.0(0.0-0.1) | 0.034 |
| T<sub>max</sub>>8s / T<sub>max</sub>>6s | 0.4(0.3-0.7) | 0.3(0.2-0.6) | 0.100 |
| T<sub>max</sub>>8s / T<sub>max</sub>>4s | 0.2(0.1-0.5) | 0.1(0.1-0.2) | 0.033 |
| T<sub>max</sub>>6s / T<sub>max</sub>>4s | 0.5(0.3-0.7) | 0.4(0.2-0.6) | 0.037 |
| Stroke etiologies |  |  | 0.619 |
| LAA | 25(73.5%) | 18(78.3%) |  |
| CE | 6(17.6%) | 2(8.7%) |  |
| Dissection | 1(2.9%) | 2(8.7%) |  |
| Undetermined | 2(5.9%) | 1(4.3%) |  |

NIHSS: National Institutes of Health Stroke Scale; ASPECTS: Alberta Stroke Program Early CT Score; T<sub>max</sub>: time-to-peak of the residual function; LAA: large-artery atherosclerosis; CE: cardioembolism;

Then, the baseline NIHSS score, ASPECTS score, and each hypoperfusion profile with P < 0.1 from the univariable analysis (i.e., the volume of T_max_>4s, T_max_>6s, T_max_>8s, T_max_>10s, and T_max_ ratios of >10s/6s, >10s/>4s, >8s/>4s, >6s/>4s) were incorporated into a separate multivariable regression model. Multivariable models indicated that the volumes of T_max_>4s (OR, 0.996; 95% CI: 0.992-1.000; P = 0.038), T_max_>6s (OR, 0.993; 95% CI: 0.988-0.999; P = 0.016), and T_max_ ratio of >6s/>4s (OR, 0.066; 95% CI: 0.005-0.874; P = 0.039) were all independent predictors of good outcome after EVT in iICAO patients after adjustment,respectively. The ROC analyses identified the optimal cutoff values, sensitivity, specificity and Youden index for the T_max_ ratio of >6s/>4s in predicting functional outcome after EVT as 0.5, 55.9%, 73.9% and 29.8%, respectively. For T_max_>4s, the values were 166.7mL, 91.2%, 47.8%, and 39.0%, while for T_max_>6s, they were 64.9mL, 82.4%, 60.9%, 43.3% (Table 4).

**Table 4.** Ability of Perfusion Lesion to Discriminate Neurological Outcome in Patients with Symptomatic Isolated Internal Carotid Artery Occlusion after Endovascular Thrombectomy T*_max_*: time-to-peak of the residual function; AUC: area under the curve.

| Variable | AUC (95% CI) | Cut off | Sensitivity | Specificity | Youden index | P value |
| --- | --- | --- | --- | --- | --- | --- |
| Baseline infarct core volume | 0.566(0.417-0.716) | 5.3mL | 44.1% | 82.6% | 26.7% | 0.384 |
| Hypoperfusion profiles |  |  |  |  |  |  |
| T<sub>max</sub>>4s, mL | 0.652(0.499-0.805) | 166.7mL | 91.2% | 47.8% | 39.0% | 0.053 |
| T<sub>max</sub>>6s, mL | 0.715(0.575-0.854) | 64.9mL | 82.4% | 60.9% | 43.3% | 0.006 |
| T<sub>max</sub>>8s, mL | 0.712(0.571-0.853) | 18.3mL | 85.3% | 56.5% | 41.8% | 0.007 |
| T<sub>max</sub>>10s, mL | 0.706(0.564-0.848) | 3.8mL | 85.3% | 56.5% | 41.8% | 0.009 |
| T<sub>max</sub>>10s / T<sub>max</sub>>8s | 0.618(0.467-0.769) | 0.7 | 26.5% | 95.2% | 21.7% | 0.146 |
| T<sub>max</sub>>10s / T<sub>max</sub>>6s | 0.645(0.498-0.793) | 0.1 | 61.8% | 68.2% | 30.0% | 0.069 |
| T<sub>max</sub>>10s / T<sub>max</sub>>4s | 0.666(0.522-0.810) | 0.0 | 79.4% | 52.2% | 31.6% | 0.034 |
| T<sub>max</sub>>8s / T<sub>max</sub>>6s | 0.631(0.483-0.779) | 0.4 | 67.6% | 59.1% | 26.7% | 0.100 |
| T<sub>max</sub>>8s / T<sub>max</sub>>4s | 0.668(0.523-0.812) | 0.1 | 82.4% | 47.8% | 30.2% | 0.033 |
| T<sub>max</sub>>6s / T<sub>max</sub>>4s | 0.664(0.522-0.806) | 0.5 | 55.9% | 73.9% | 29.8% | 0.037 |

## Discussion

In this multicenter real-world study, we found that a lower T_max_ ratio of >6s/>4s was an independent predictor for a higher risk of neurological deterioration before EVT, and it was independently associated with a better functional outcome after EVT for AIS patients with iICAO. A threshold of T_max_ >6s/>4s of 0.5 enabled prediction of neurological deterioration and functional outcome.

The optimal treatment strategies for iICAO patients still remain uncertain and depend on the expertise. Previous studies have provided limited data regarding EVT for acute symptomatic iICAO. According to the current AHA/ASA guideline based on large EVT clinical trials, EVT is a well-proven treatment for LVO including distal ICA and MCA^5^. A limited number of AIS patients with iICAO were included in those clinical trials. Some researches with small samples have indicated that EVT might be an alternative approach for symptomatic iICAO patients by mitigating the risk of neurological deterioration secondary to hemodynamic insufficiency and subsequent infarction^7,17^. A retrospective observational study from MR CLEAN analyzed a total of 51 iICAO patients and found that those who underwent EVT demonstrated better outcome compared to those receiving medical treatment alone^6^. Recently, a retrospective analysis of 45 iICAO patients from ETIS registry suggested that EVT for AIS (<2weeks) due to iICAO was associated with a high recanalization rate and a 40% rate of good functional outcome^7^. These results were comparable to those observed in AIS patients with intracranial occlusions following EVT. Nearly half of AIS patients undergoing EVT did not have good outcome despite successful recanalization, a phenomenon referred to as futile recanalization^18^. This suggested that successful recanalization might not necessarily correspond to effective reperfusion of ischemic brain tissues. The inadequate perfusion status of brain tissue would be related with futile recanalization due to the no-reflow phenomenon or reperfusion injury^19,20^.

It is worth exploring the role of perfusion status before EVT in facilitating appropriate triage of iICAO patients for EVT. Brain tissue-level perfusion parameters on perfusion imaging reflected the comprehensive manifestation of the arterial collateral blood flow successfully permeated to the brain tissue and then transit to venous circulation. CTP could provide the quantitative information of cerebral perfusion, which might contribute to the selection of the optimal candidates for reperfusion therapy. The perfusion pattern in patients with iICAO characterized by a small core but a large hypoperfusion might be differ from perfusion profiles seen in AIS patients with intracranial occlusions. Good collateral circulation via the circle of Willis could maintain perfusion for the brain tissue in the affected hemisphere^21^. The neurological deficits would be exacerbated when hypoperfusion prolonged as a result of inadequate treatment and collateral circulation failure^22^. The iICAO patients with neurological symptoms implying hypoperfusion might due to collapsed collateral circulation. Therefore, tissue-level perfusion status measured with CTP might be helpful in refining the selection of EVT-eligible iICAO patients.

T_max_ mismatch ratios offer a comprehensive evaluation of perfusion delay status of brain tissue, which might be useful for selection of iICAO patients eligible for endovascular intervention. For AIS patients with intracranial LVO, previous studies showed that the ratio of T_max_>10s />6s was associated with fast infarct progression and functional outcome^12,14^. And a lower T_max_ ratio of >10 s/>6 s was found to be associated with intracranial atherosclerotic LVO characterized as chronic hypoperfusion^14,23^. A previous study has demonstrated that a profile of T_max_ >4 s/ >6s with ≥2 was an independent indicator of underlying intracranial atherosclerotic disease^24^. The threshold of T_max_ >4 s has been defined as brain tissues likely exhibiting benign oligemic, whereas T_max_ >6 s signifies brain tissues at risk, commonly referred to as penumbral lesions. T_max_ >10 s is indicative of brain tissues experiencing malignant hypoperfusion^14,23,24^. For patients with iICAO, the perfusion status might be different from that observed in AIS patients with intracranial LVO. In our study, we observed that a perfusion profile characterized by a lower T_max_ >6s/>4s was independently associated with a higher rate of neurological deterioration and better functional outcome after EVT in iICAO patients, suggesting that a cutoff value of 0.5 might be optimal for EVT. Consequently, our findings might reflect the presence of clinically silent oligemic brain tissues and penumbra lesions that are at high risk for progression. However, these hypoperfused brain tissues could still be salvaged through EVT intervention, resulting in functional recovery.

There are several limitations of this study. Firstly, as this study was based on retrospective analyses, our findings might have selection bias and require prospective validation. Secondly, this multicenter study consisting of a selected population in which eligibility for EVT was decided by the local stroke teams, and lacked data from patients who did not receiving EVT were lacking. Lastly, the sample size of this study was relatively small. Future investigations into the outcomes following EVT in iICAO patients should be conducted through larger sample sizes and randomized prospective clinical trials.

## Conclusion

In summary, our findings have suggested perfusion defect severity ratio of T_max_ >6s/>4s less than 0.5 might indicate an elevated risk of neurological deterioration, yet it would be associated with achieving better outcome after EVT for iICAO patients. Therefore, the hypoperfusion profile characterized by a T_max_ >6s/>4s on CTP could be valuable for the neurointerventionists in facilitating appropriate patient triage and guiding prognosis for EVT in iICAO patients.

## Abbreviations

iICAO: isolated internal carotid artery occlusion
AIS: acute ischemic stroke
NIHSS: National Institutes of Health Stroke Scale
OPT: onset-to-puncture time
HT: hemorrhagic transformation
EVT: endovascular treatment
LVO: large vessel occlusion
CTA: CT angiography
CTP: CT perfusion
ROC: receiver operating characteristic
AUC: area under the curve
mRS: modified Rankin Scale score
LAA: large-artery atherosclerosis
CE: cardioembolism
TICI: Thrombolysis in Cerebral Infarction
CBF: cerebral blood flow
T*_max_*: time-to-peak of the residual function

## Disclosure

None declared.

## Data availability statement

The data that support the findings of this study are available from the corresponding author upon reasonable request.

## Sources of Funding

This work was support by the National Natural Science Foundation of China (82402210), Natural Science Foundation of Zhejiang Province (LQ23H090014) and Medical Health Science and Technology Program of Zhejiang Province (2023KY778).

## Notes

### Competing Interest Statement

The authors have declared no competing interest.

### Author Declarations

the local human ethics committees of Sir Run Run Shaw Hospital of Zhejiang University, School of Medicine

